# GLP-1 RA-Exacerbated Gut Microbiome Dysbiosis in Obesity Mediates Post-Cessation Weight Regain

**DOI:** 10.64898/2026.05.02.26352300

**Authors:** Delei Song, Yuhang Ma, Yi Lin, Yulong Han, Zhiyi Wang, Zhichao Feng, Yongde Peng, Yu Shi, Baohai Pan, Feng Zhang, Rui Zhai, Ying Zhu, Huize Miao, Xiaoying Ding, Chenhong Zhang

## Abstract

GLP-1 receptor agonists (GLP-1 RAs) effectively reduce weight in obesity, although significant weight regain typically follows discontinuation. Here, in a randomized clinical trial (ChiCTR2200066014), we found that GLP-1 RA (semaglutide) and a high-fibre diet achieved similar 12-week weight reduction, but semaglutide recipients exhibited significantly higher weight rebound at the 14^th^ week after intervention cessation. Shotgun metagenomic sequencing revealed that semaglutide aggravated the proinflammatory signature in the gut microbiome, which contrasted with high-fibre diet intervention. The microbiota transplanted from semaglutide-treated subjects to germ-free mice induced gut barrier dysfunction, systemic inflammation and an increase in the bacterial antigen load in the liver and adipose tissue, which activated the NF-κB pathway to drive lipid accumulation. Using a diet-induced obesity mouse model, we found that semaglutide exacerbated gut microbiome dysbiosis by weakening host immune surveillance of the gut microbiota through downregulating IFN-γ to reduce antimicrobial peptides expression and delaying gut transit time to shift microbial metabolism from saccharolysis towards proteolysis. Crucially, combining semaglutide with dietary fibre in mice mitigated microbiome dysbiosis and attenuated weight regain post-cessation. These findings suggest that GLP-1 RA-exacerbated gut microbiome dysbiosis in obesity as a key mediator of post-treatment weight rebound and propose adjunctive fibre supplementation as a strategy to sustain weight loss.

## Introduction

As obesity is increasingly recognized as a complex chronic disease, advanced treatments such as glucagon-like peptide-1 receptor agonists (GLP-1 RA, e.g., liraglutide, semaglutide, and tirzepatide) have emerged^1,2^. These agents reduce weight by 6.0%-17.8% within 52 to 72 weeks of monotherapy while improving glucose control and reducing the risks of cardiovascular and liver diseases^3–6^. Their mechanisms involve enhancing satiety, reducing chronic inflammation, delaying gastric emptying, and modulating energy balance^7–9^.

However, weight regain after cessation is a critical limitation. Clinical data demonstrate rebound, such as recovery of two-thirds of lost weight within one year after semaglutide cessation or 82.5% of participants regaining more than 25% of the lost weight in one year after tirzepatide cessation^10–12^. A meta-analysis with 37 studies showed that the average estimated weight regain was 9.9 kg within the first year after semaglutide and tirzepatide cessation, with a projected return to baseline weight at an average of 1.5 years^13^. Notably, weight regain following treatment cessation is significantly correlated with the extent of reversal in cardiometabolic improvements^12^. This rebound is mechanistically linked to physiological reversals, including rebound increases in hunger hormone (e.g., ghrelin), declines in satiety hormones, reduced energy expenditure and increased fat storage, although precise pathways remain incompletely defined^14,15^. Sustained treatment with these GLP-1 RAs is necessary for maintained efficacy^16^, and these agents have been approved for chronic weight management in an increasing number of countries, yet economic burdens and long-term safety require evaluation. Understanding rebound mechanisms is thus pivotal for optimizing obesity management with GLP-1 RAs.

Dysbiosis of the gut microbiome plays a causal role in the development of obesity. The mechanisms involve specific members of gut microbiota-derived metabolites (e.g., shorty-chain fatty acids, bile acids and indole derivatives) and other active molecules (e.g., lipopolysaccharides) modulating energy metabolism, inflammation pathways and the appetite-regulating neural system^17^. Then, the gut microbiome serves as a pivotal therapeutic target for obesity management. Lifestyle interventions, especially dietary modifications, promote weight loss by beneficially regulating the composition and function of the gut microbiota^18^.

Previous studies have shown that GLP-1 RAs can change the gut microbiota, yet inconsistencies have been reported. Some studies have observed increased beneficial bacteria (e.g., *Akkermansia muciniphila*)^19,20^, while others have noted elevated opportunistic pathogens (e.g., Enterobacteriaceae spp.)^21,22^. Critically, clinical trial data on GLP-1 RA-induced microbiota alteration remain sparse, with functional impacts unclear. A previous study demonstrated that the GLP-1 receptor of gut intraepithelial lymphocytes (IELs) controls a subset of GLP-1 RA actions on gut microbiota, but the effect of GLP-1 RA on gut microbiota may operate through multifactorial mechanisms and intertwine with GLP-1 RA’s role in improving host metabolism^21^. Notably, GLP-1 RA-modified gut microbiota would persist post-cessation, potentially influencing weight rebound. Thus, alterations in gut microbiota structure and function during GLP-1 RA treatment and the impact of such changes after cessation require rigorous investigation.

In the current randomized, multicenter clinical study, we showed that semaglutide treatment induces a proinflammatory gut microbiome signature distinct from high-fibre diet intervention, despite comparable short-term weight loss in obese and overweight subjects. However, the subjects with semaglutide treatment had higher weight regain after 14 weeks of cessation. By fecal microbiome transplantation, we showed that semaglutide-exacerbated gut microbiome dysbiosis induced impaction of the gut barrier, chronic low-grade systemic inflammation and acceleration of lipid accumulation in the liver and adipose tissue of germ-free mice. In a diet-induced obesity (DIO) mouse model, we showed that a decrease in antimicrobial peptides (AMPs) induced by modulation of the IFN-γ/STAT3 pathway and a longer gut transit time associated with a shift in microbial metabolism from saccharolysis towards proteolysis contributed to the actions of GLP-1 RA on the gut microbiota. We also proved that combination with dietary fibre supplementation could mitigate adverse effects of GLP-1 RA on gut microbiota and attenuate weight regain following treatment cessation.

## Results

### GLP-1 RA recipients had higher weight rebound than dietary fibre recipients following intervention cessation

From 18 December 2022 to 13 March 2023, a total of 83 participants were randomly assigned to receive once-weekly subcutaneous semaglutide (G group, *n*=34, 0.25 mg for weeks 1-2, 0.5 mg for weeks 3-4, 1.0 mg for week 5 onwards) or high-fibre diet intervention (W group, *n*=49, intake of 45 g dietary fibre per day, see methods and Figure 1a; Table S1). At baseline, the body weight, waist circumference, waist-to-hip ratio (WHR), body fat mass, fasting C-peptide, serum creatinine, and anxiety score in the G group were significantly higher than those in the W group. After adjusting for body weight, there were no significant differences in the aforementioned parameters between the G and W groups (Table S2). On the other hand, there was no difference between the two groups in total intake of energy, fat, protein, carbohydrate or fibre (Table S3).

**Figure 1.**
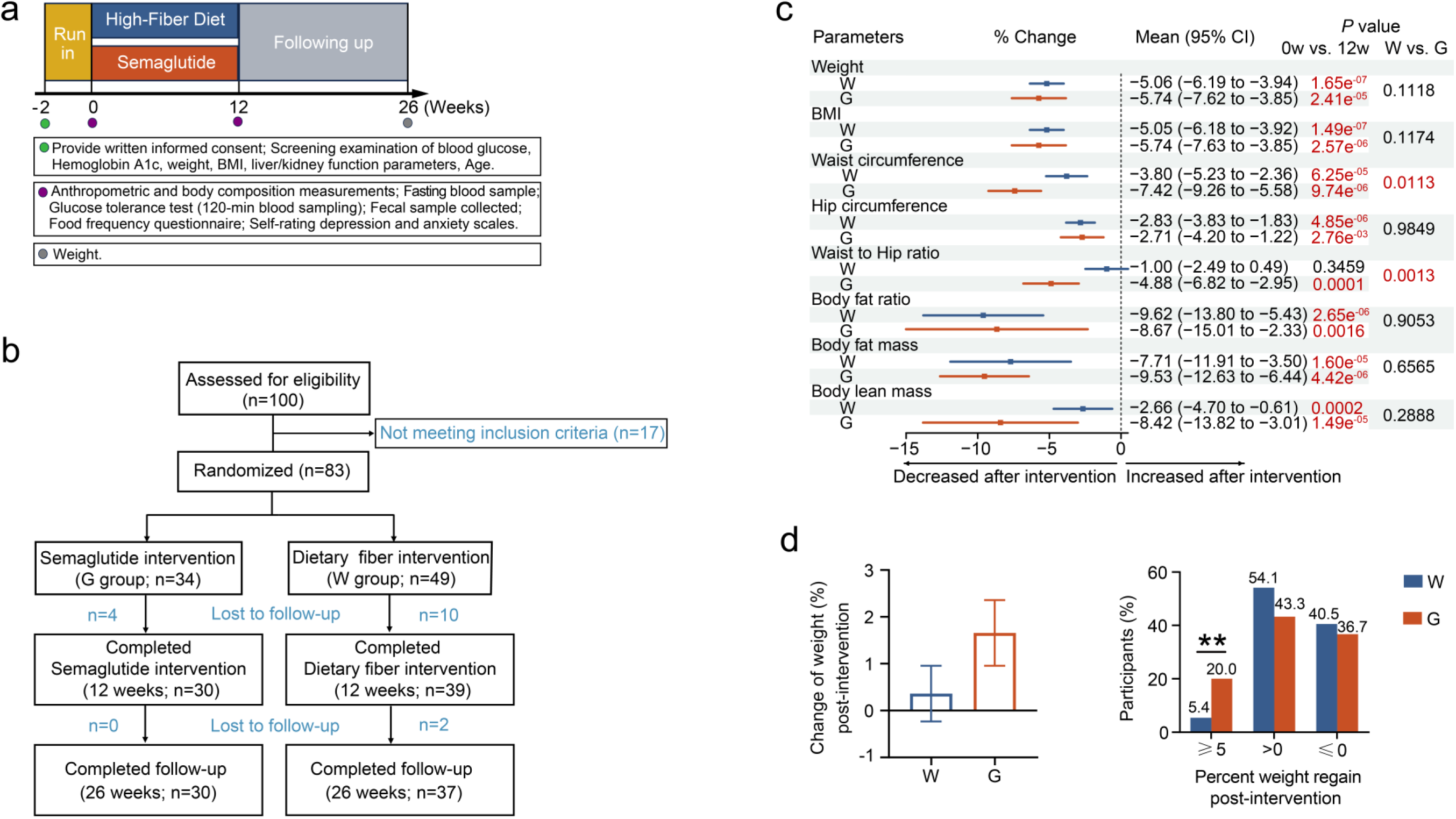
GLP-1 RA recipients had higher posttreatment weight rebound than fibre recipients. **a**, Clinical study design. **b**, Flow diagram of subject enrollment, intervention allocation, and follow-up procedures. **c**, Percentage changes in anthropometric and body composition parameters in the G (*n*=30) and W (*n*=39) groups post-intervention. The square within the middle of the 95% confidence interval (CI) represents the mean value. **d**, Weight regain in the G (*n*=30) and W (*n*=37) groups at the 14^th^ week post-intervention. Data are presented as the mean ± s.e.m. In **a** and **c**, BMI, body mass index. In **c** and **d**, inter-group comparisons at the same timepoint used a two-tailed Mann‒Whitney U test; intra-group comparisons at different timepoints used a two-tailed Wilcoxon matched-pair signed-rank test; comparisons of percentage weight regain between groups used Pearson’s chi-squared test. \*\**P* < 0.01.

Overall, 69 participants (83.1%) completed the 12-week intervention (30 participants in the G group and 39 participants in the W group; Figure 1b). No serious adverse events were reported during the trial. Compared to the baseline, the participants in the G group had significantly decreased intake of total energy and all macronutrients during the intervention, except for no change in intake of protein (Table S3). However, the intake of energy and all the macronutrients of the participants in the W group showed no significant changes, except for a significant increase in fibre intake by design (Table S3).

As the primary outcome, the mean percentage change in body weight from baseline to 12 weeks was -5.06% (95% confidence interval [CI], -6.19 to -3.94) in the W group and -5.74% (95% CI, -7.62 to -3.85) in the G group (Figure 1c; Table S4).

When comparing the clinical parameters between the two groups after intervention, the baseline body weight was adjusted. There was no significant difference between the two groups in the change in body weight (Figure 1c; Table S4). Moreover, the participants in the two groups had significant reductions from baseline in BMI, waist and hip circumferences, while the G group had higher decreases in waist circumference and WHR than the W group (Figure 1c; Table S4). The body fat mass at 12 weeks was significantly reduced from the baseline in both the G and W groups, with no significant difference between the groups (Figure 1c; Table S4). Although both the semaglutide and high-fibre diet intervention led to loss of body lean mass, the reduction was less in the W group than in the G group (Table S5).

Both 12 weeks of semaglutide and high-fibre diet intervention led to a reduction in 2 h postprandial plasma glucose, and the change in the G group was higher than that in the W group; fasting insulin, 2 h postprandial plasma insulin and homeostasis model assessment 2 of insulin resistance (HOMA2-IR) were only significantly decreased in the W group, while only semaglutide treatment increased fasting C-peptide but increased HOMA2-IR (Table S5). For blood lipids, LDL cholesterol and triglycerides were significantly reduced from baseline in the W group, and HDL cholesterol was significantly decreased in the G group (Table S5). Moreover, both the G and W groups had significant improvements in liver and kidney function (Table S5).

To evaluate the regain of body weight after withdrawal of semaglutide or high-fibre diet intervention, we conducted a follow-up with our participants at the 14^th^ week post-intervention. Thirty-seven subjects in the W group and 30 subjects in the G group took part in the assessment of their body weight. There was no significant difference in the change in body weight between the two groups, but subjects in the W group tended to show less weight regain than those in the G group (Figure 1d). Notably, the percentage of participants with a body weight regain of more than 5% was much higher in the G group than in the W group (20.0% in the G group vs. 5.4% in the W group, *P*=0.0075; Figure 1d).

Taken together, 12-week intervention with semaglutide and a high-fibre diet induced similar decreases in body weight in subjects with obesity, and the high-fibre diet resulted in smaller reductions in lean body mass and improved insulin resistance. More importantly, semaglutide recipients exhibited much higher weight rebound than dietary fibre recipients following intervention cessation.

### Gut microbiota dysbiosis is alleviated by high-fibre diet intervention, but exacerbated by GLP-1 RA treatment

We performed shotgun metagenomic sequencing of the gut microbiota of the participants in both groups at baseline and at the end of the 12-week treatment. A total of 3692 non-redundant high-quality metagenome-assembled genomes (HQMAGs) were de novo reconstructed, representing over 89.1% of the total reads from 138 fecal samples. We calculated the proportion of gut microbiota variance explained by potential influencing factors, among which the individual variability explained most of the variation in microbiota composition (Figure S1a). Therefore, subject was regarded as a covariate for adjustment in subsequent analyses. At baseline, there was no difference in the diversity and whole structure of the gut microbiota in obese participants between the G and W group (Figure 2a and 2b). After 12 weeks of intervention, a significant increase in the richness and diversity of the gut microbiota was observed in the G group but not in the W group (Figure 2a). Subject-adjusted principal coordinate analysis (aPCoA) of Bray‒Curtis distance and permutational multivariate analysis of variance (PERMANOVA; 999 permutations) based on the abundances of HQMAGs demonstrated that the whole structure of gut microbiota showed significant alterations in both the G and W groups compared to baseline (Figure 2b). Then, the participants with semaglutide or a high-fibre diet had significantly distinguished gut microbiota at the end of the intervention (Figure 2b), although the weight loss was comparable between interventions.

**Figure 2.**
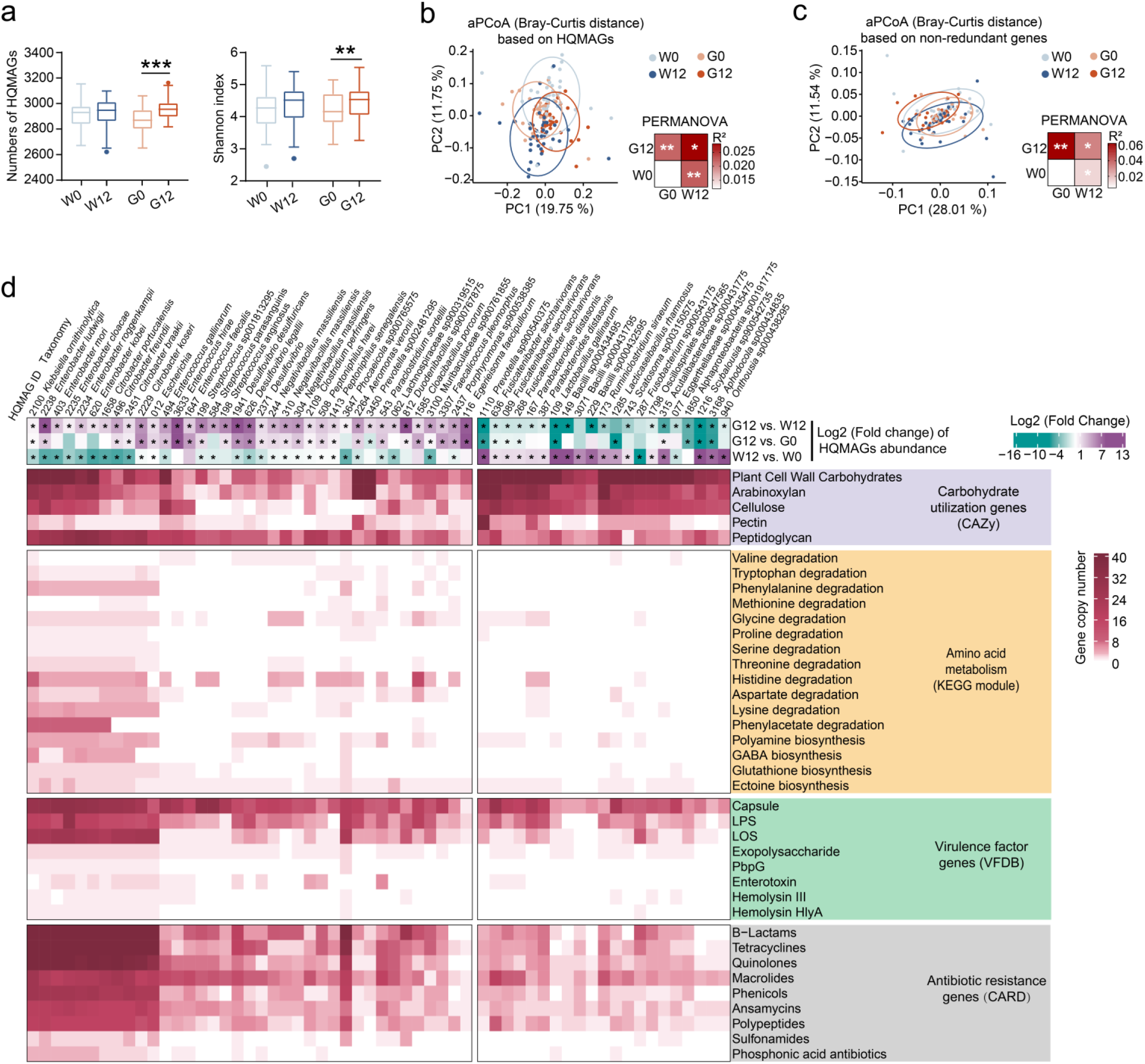
Gut microbiota dysbiosis in obese subjects is exacerbated by GLP-1 RA treatment but alleviated by high-fibre diet intervention. **a**, α-diversity of the gut microbiome. Boxplots: centerline, median value; box, first and third quartiles; whiskers, 1.5× interquartile range with outliers individually plotted. **b** and **c**, Adjusted principal coordinates analysis (aPCoA) score plot based on Bray‒Curtis distance at high-quality metagenome-assembled genomes (HQMAGs; **b**) and non-redundant genes (**c**) levels during intervention. Statistical analyses were performed using permutational multivariate analysis of variance (PERMANOVA; 999 permutations), \**P* < 0.05, \*\**P* < 0.01. Subject adjusted in both aPCoA and PERMANOVA. The circle represents the 95% confidence interval. **d**, Gene copy numbers carried by the HQMAGs with abundance differences between the G and W groups. Only the genes with significant differences (*P* < 0.05) between groups are presented. Asterisks indicate significant differences (*P* < 0.05) in HQMAG abundances within or between groups. In **a** and **d**, inter-group comparisons at the same timepoint used a two-tailed Mann‒Whitney U test; intra-group comparisons at different timepoints used a two-tailed Wilcoxon matched-pair signed-rank test. \*\**P* < 0.01, \*\*\**P* < 0.001. In all panels, *n*=30 in the G group, *n*=39 in the W group. G0, baseline of the G group. G12, post-intervention G group. W0, baseline of the W group. W12, post-intervention W group.

Using the Boruta model and Wilcoxon matched-pair signed-rank test, we found that the abundances of opportunistic pathogens, such as HQMAGs of *Enterococcus faecalis*, *Enterococcus gallinarum Enterobacter ludwigii*, *Citrobacter koseri*, *Streptococcus anginosus*, and *Clostridium perfringens*, significantly increased in the G group after intervention; in contrast, the abundance of opportunistic pathogens significantly decreased in the W group after intervention, while the abundances of potentially beneficial bacteria, such as HQMAGs of *Parabacteroides distasonis* and *Fusicatenibacter saccharivorans*, significantly increased (Figure S1b; Table S6). These alterations led to a significantly higher abundance of opportunistic pathogens in the G group than in the W group at the end of the intervention, whereas the abundance of potential beneficial bacteria in the W group was significantly higher than that in the G group (Figure S1b; Table S6).

The aPCoA of Bray‒Curtis distance based on the abundance of non-redundant genes revealed that the gut microbiome functions showed significant alterations in both the G and W groups compared to baseline (statistical analyses by PERMANOVA with 999 permutations; Figure 2c). To investigate the specific functional changes, we annotated the non-redundant genes using the Kyoto Encyclopedia of Genes and Genomes (KEGG) database, carbohydrate-active enzymes database (CAZy), virulence factors database (VFDB), and comprehensive antibiotic resistance database (CARD). Compared with baseline, the abundances of genes for branched-chain amino acids, basic amino acids, and acidic amino acids degradation significantly increased in the G group post-intervention (Figure S1c). Moreover, the abundances of mucin utilization genes significantly increased, whereas the abundances of genes for plant cell wall carbohydrates, pectin, arabinoxylan, and cellulose utilization significantly decreased in the G group post-intervention (Figure S1d). We further found that post-intervention, virulence factor genes (VFGs) involved in bacterial adhesion, biofilm formation, biosynthesis of bacterial antigens capable of inducing host inflammatory responses, and antibiotic resistance genes (ARGs) showed significantly increased abundances in the G group (Figure S1e and S1f). In the W group, there were no significant changes in the abundances of branched-chain amino acids and acidic amino acids degradation genes, arabinoxylan and cellulose utilization genes, or biofilm formation VFGs post-intervention (Figure S1c and S1d and S1f). Conversely, the abundance changes of histidine degradation genes and lipopolysaccharides (LPS) biosynthesis genes with a significant decrease post-intervention in the W group exhibited an opposite pattern to that in the G group (Figure S1c and S1e). Although the post-intervention patterns of abundance changes in the genes responsible for the utilization of mucin, plant cell wall carbohydrates, and pectin, VFGs involved in bacterial adhesion, biosynthesis of bacterial antigens, and ARGs in the W group were opposite to those in the G group, the specific genes that changed were different from those in the G group (Figure S1d-S1f). Additionally, we observed that in the W group, the abundances of aromatic amino acid degradation genes and VFGs involved in bacterial invasion decreased significantly, while the abundance of starch utilization genes increased significantly post-intervention (Figure S1c and S1d and S1e).

The overall structures of the gut microbiota shaped by semaglutide and high-fibre diet interventions were different; thus, the functional profiles of the microbiota shaped by the two types of intervention were distinguished significantly (Figure 2c). Compared with the W group, the HQMAGs with higher abundance in the G group carried significantly more copies of genes involved in amino acid degradation, biosynthesis of bacterial antigens capable of inducing host inflammatory responses, and antibiotic resistance. In contrast, the HQMAGs with higher abundance in the W group had significantly more copies of genes involved in carbohydrates utilization (Figure 2d).

Taken together, the metagenomic data from our above clinical trial showed that a 12-week intervention with semaglutide exacerbated gut microbiota dysbiosis by enriching opportunistic pathogens and induced attenuated capability of carbohydrate metabolism alongside enhanced proteolytic-related pathways in the gut microbiome of the participants.

### Gut microbiota in GLP-1 RA recipients induced chronic inflammation and led to lipid accumulation

We transplanted the participants’ fecal microbiota collected at the end of 12 weeks of semaglutide or high-fibre diet intervention to germ-free mice by oral gavage 3 times a week. The animal trial was repeated in two independent trials, and the results of the two independent trials were consistent. PCoA based on 16S rRNA gene V3-V4 region sequencing revealed that the structure of the gut microbiota in the mice with a microbiome from semaglutide recipients (M_FG_) was significantly different from that in the mice that received a microbiome from subjects in the high-fibre diet group (M_FW_; Figure S2a and S2b). The gut microbiome of the recipient mice was more similar to that of their corresponding human donors. Specifically, the M_FG_ mice had higher opportunistic pathogenic *Enterobacter* spp. and *Citrobacter* spp., while the M_FW_ mice had much more *Parabacteroides* spp. and *Fusicatenibacter* sp. (Figure S2c; Table S7).

After two weeks of fecal microbiota transplantation, the M_FG_ mice had significantly higher weight and increased adipocyte size in the epididymal and subcutaneous fat pad than the M_FW_ mice, although there was no significant difference in body weight or food intake (Figure 3a and 3b; Figure S3a-S3d). The liver non-alcoholic fatty liver disease (NAFLD) score from hematoxylin‒eosin (H&E) staining and liver lipid content from Oil Red O staining were also significantly higher in the M_FG_ group than in the M_FW_ group (Figure 3c; Figure S3e). Moreover, the levels of triglycerides were significantly increased in both the liver and epididymal fat of M_FG_ mice (Figure 3d; Figure S3f). Taken together, the microbiome from subjects treated with semaglutide accelerated lipid accumulation in the liver and adipose tissue in mice, suggesting that the GLP-1 RA-shaped microbiome contributes to inducing the regain of body weight in obese people after withdrawal of treatment.

**Figure 3.**
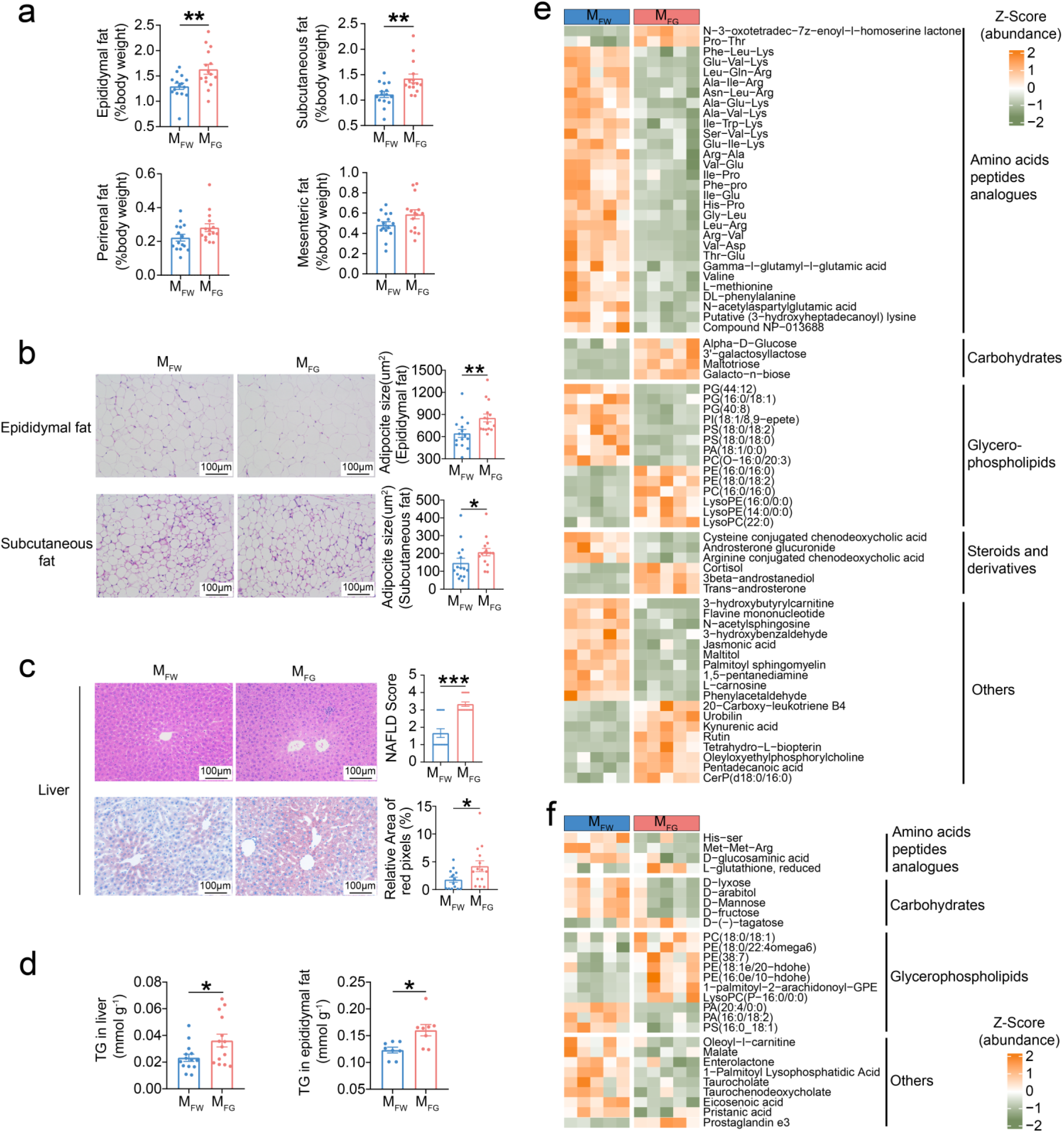
Gut microbiota in GLP-1 RA recipients led to lipid accumulation in germ-free mice. **a**, Weight coefficients of adipose tissues. **b**, Representative H&E-stained sections of epididymal and subcutaneous fats (scale bar = 100 μm) and calculated mean adipocyte area. **c**, Representative H&E-stained and Oil red o-stained liver sections (scale bar = 100 μm) and calculated non-alcoholic fatty liver disease (NAFLD) score and relative area of red pixels. **d**, Triglyceride (TG) contents in liver (*n*=15 per group) and epididymal fat (*n*=8 per group). **e** and **f**, Heatmaps show the abundances (Z scored) of differentially abundant metabolites of cecal content (**e**) and serum (**f**) between groups (*n*=5 per group). In **a** to **c**, *n*=15 per group. In **a** to **d**, data after batch effect correction using the ComBat method and presented as the mean ± s.e.m. In all panels, statistical analyses were performed using unpaired two-tailed Student’s *t* test (**a**, **e**, **f**) or two-tailed Mann‒Whitney U test (**b**, **c**, **d**). \**P* < 0.05, \*\**P* < 0.01, \*\*\**P* < 0.001. M_FW_, mice received microbiome from high-fibre diet recipients. M_FG_, mice received microbiome from semaglutide recipients.

We compared the fecal and serum metabonomic profiles of the above gnotobiotic mice by LC‒MS, and orthogonal partial least squares discriminant analysis (OPLS-DA) showed significant differences in both fecal and serum metabolites between M_FG_ and M_FW_ mice (Figure S4a and S4b). Compared to the M_FG_ mice, the M_FW_ mice exhibited lower levels of oligosaccharides (galacto-n-biose, galactosyllactose, and maltotriose) in the gut, alongside higher concentrations of monosaccharides (mannose, fructose, and lyxose) in the blood (Figure 3e and 3f; Figure S4c and S4d; Table S8). This was consistent with the enhanced carbohydrate metabolism capability of the gut microbiota observed in subjects receiving dietary fibre intervention. Notably, compared to the M_FW_ mice, the M_FG_ mice had lower levels of oligopeptides in the gut and blood (Figure 3e and 3f), which was consistent with the enhanced proteolytic capability of the gut microbiota observed in subjects receiving semaglutide intervention. Additionally, many kinds of metabolites increased in the gut or blood of M_FG_ mice, such as lyso-phosphatidylethanolamine, phosphatidylethanolamine and N-3-oxotetradec-7z-enoyl-l-homoserine lactone (quorum sensing signal of gram-negative bacteria), may be associated with a damaged gut barrier and elevated inflammation levels in these mice compared to the M_FW_ group (Figure 3e and 3f; Figure S4c and S4d; Table S8).

By RT‒qPCR, we found that the gene expression of proinflammatory cytokines, such as IL-1β, IL-6 and TNF-α, was significantly increased in the colon of the M_FG_ compared with that of the M_FW_ mice, suggesting that the microbiome from semaglutide-treated subjects induces higher inflammation in mice (Figure 4a; Figure S5 and S6). Moreover, immunohistochemical detection of tight junction-related proteins and mucin in colonic tissue showed that the M_FG_ mice had lower levels of ZO-1 and MUC2 (Figure 4b; Figure S5). We also found that lipopolysaccharides-binding protein (LBP), which can bind to antigens such as endotoxin produced by bacteria and represents a surrogate biomarker that links bacterial antigen load in the blood with host inflammatory response, was significantly increased in the serum of M_FG_ mice compared with M_FW_ mice by using ELISA (Figure 4c). Subsequently, we measured inflammation-related cytokines in serum by mesoscale discovery-electrochemiluminescence and found that the levels of proinflammatory cytokines, such as IL-1β, IL-6, TNF-α and IFN-γ, were significantly higher in M_FG_ mice (Figure 4c; Figure S5 and S6).

**Figure 4.**
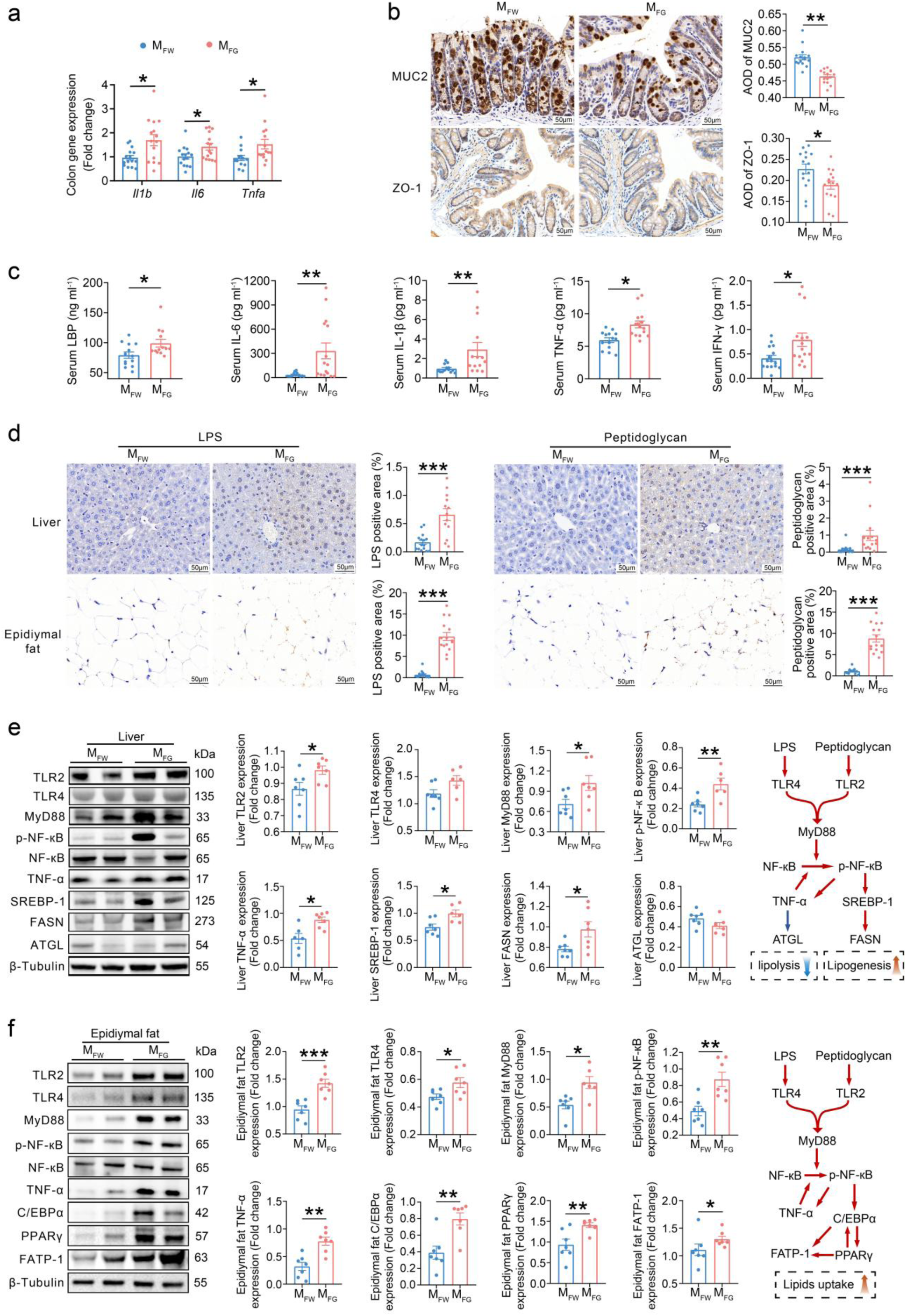
Gut microbiota in GLP-1 RA recipients induces gut barrier dysfunction, activates the TLRs/MyD88/NF-κB inflammatory pathway, drives lipogenesis and suppresses lipolysis in mice. **a**, Relative expression of *Il1b*, *Il6* and *Tnfa* in the colon measured by RT‒qPCR. **b**, Representative images of MUC2 and ZO-1 immunohistochemical staining in the colon (scale bar = 50 μm) and the calculated average optical density (AOD). **c**, Serum lipopolysaccharide binding protein (LBP) level measured by ELISA, and serum proinflammatory cytokines levels measured by meso scale discovery-electrochemiluminescence. **d**, Representative images of lipopolysaccharide (LPS) and peptidoglycan immunohistochemical staining in liver and epididymal fat (scale bar = 50 μm) and calculation of the positive areas. **e** and **f**, Representative western blot images and quantitative analysis of key regulators in the TLRs/MyD88/NF-κB inflammatory pathway and lipid metabolism in liver (**e**) and epididymal fat (**f**). *n*=7 per group. The flow chart shows a schematic representation of the TLRs/MyD88/NF-κB signalling pathway and its regulatory effects on lipid metabolism. Red and blue arrows represent activation and inhibitory effects, respectively. In **a** to **d**, *n*=15 per group. In all panels, data after batch effect correction using the ComBat method are presented as the mean ± s.e.m. Statistical analyses were performed using unpaired two-tailed Student’s *t* test (**a**, **e**, **f**) or two-tailed Mann‒Whitney U test (**b**, **c**, **d**). \**P* < 0.05, \*\**P* < 0.01, \*\*\**P* < 0.001. M_FW_, mice received microbiome from high-fibre diet recipients. M_FG_, mice received microbiome from semaglutide recipients.

Next, we used immunohistochemical staining to show significantly elevated LPS and peptidoglycan accumulation in the liver and adipose tissue of M_FG_ mice compared to M_FW_ mice (Figure 4d; Figure S5). By western blotting, we also found that the expression of toll-like receptor 4 (TLR4) and TLR2, which recognize bacterial antigens, and their intracellular core adaptor protein, myeloid differentiation primary response 88 (MyD88), was markedly upregulated in the liver and adipose tissue of M_FG_ mice (Figure 4e and 4f; Figure S5). In both the liver and adipose tissue of M_FG_ mice, nuclear translocation of nuclear factor kappa B (NF-κB) was significantly increased and enhanced the levels of TNF-α, suggesting the activation of the NF-κB pathway and induction of inflammation. In the liver of M_FG_ mice, NF-κB upregulated the downstream transcription factor sterol regulatory element-binding protein 1 (SREBP-1), thereby enhancing the expression of the key enzyme fatty acid synthase (FASN) in the lipogenesis pathway. Meanwhile, the level of the key lipolysis enzyme adipose triglyceride lipase (ATGL) was downregulated (Figure 4e; Figure S5 and S6). In the adipose tissue of M_FG_ mice, CCAAT/enhancer-binding protein α (C/EBPα) and peroxisome proliferator-activated receptor γ (PPARγ) were upregulated, thereby increasing the level of fatty acid transport protein 1 (FATP-1), which facilitated the uptake of long-chain fatty acids into cells (Figure 4f; Figure S5 and S6). These results indicate that compared to that in M_FW_ mice, the activation of NF-κB signalling in M_FG_ mice induced enhanced lipogenesis and reduced lipolysis in the liver and increased lipid uptake in adipose tissue.

Taken together, the gut microbiome shaped by GLP-1 RA treatment compromised the gut barrier and increased bacterial antigen loading in the host’s systemic circulation, leading to chronic low-grade inflammation, which consequently accelerated lipid accumulation in the liver and adipose tissue.

### GLP-1 RA exacerbated gut microbiome dysbiosis through a decrease in AMPs and disruption of intestinal nutrition

Since the gut microbiome shaped by GLP-1 RA treatment plays an important role in the regain of body weight after treatment cessation, we investigated the mechanism through which GLP-1 RA modulates the gut microbiome. We fed C57BL/6J mice (male, 8 weeks) a Western-style diet for 4 weeks to construct a DIO model and then performed subcutaneous injection once every three days with semaglutide (0.1 mg/kg body weight, M_G_ group) or normal saline as a control (M_C_ group) for 4 weeks. As anticipated, semaglutide treatment significantly decreased body weight, amounts of epididymal, perirenal and mesenteric fat, and amounts of fat in the liver compared to those in the control group (Figure 5a-5d). Then, we analysed semaglutide-induced gut microbiome alterations in DIO mice. By qPCR of the 16S rRNA gene, we found that the bacterial loading was significantly higher in the M_G_ group (Figure S7a). By fecal 16S rRNA gene sequencing, we showed that the richness of gut microbiota (observed ASVs) was significantly higher in the M_G_ group (Figure S7b). Moreover, a PCoA score plot based on Bray‒Curtis distance showed distinguished gut microbiota structures between the M_G_ and M_C_ groups (Figure S7c). We also found that bacteria from *Escherichia−Shigella, Alistipes, Peptococcus, Enterococcus,* and *Clostridium* were significantly enriched in the M_G_ group (Figure S7d; Table S9). Similar to our results from the clinical trial, these results from DIO mice suggested that concomitant with obesity amelioration, treatment with semaglutide induced higher microbial cell density, diversity, and abundance of opportunistic pathogens. Notably, we found that the levels of TNF-α and IFN-γ in serum were significantly decreased during drug administration (Figure S7e), which suggested that systemic inflammation was attenuated by drug administration.

**Figure 5.**
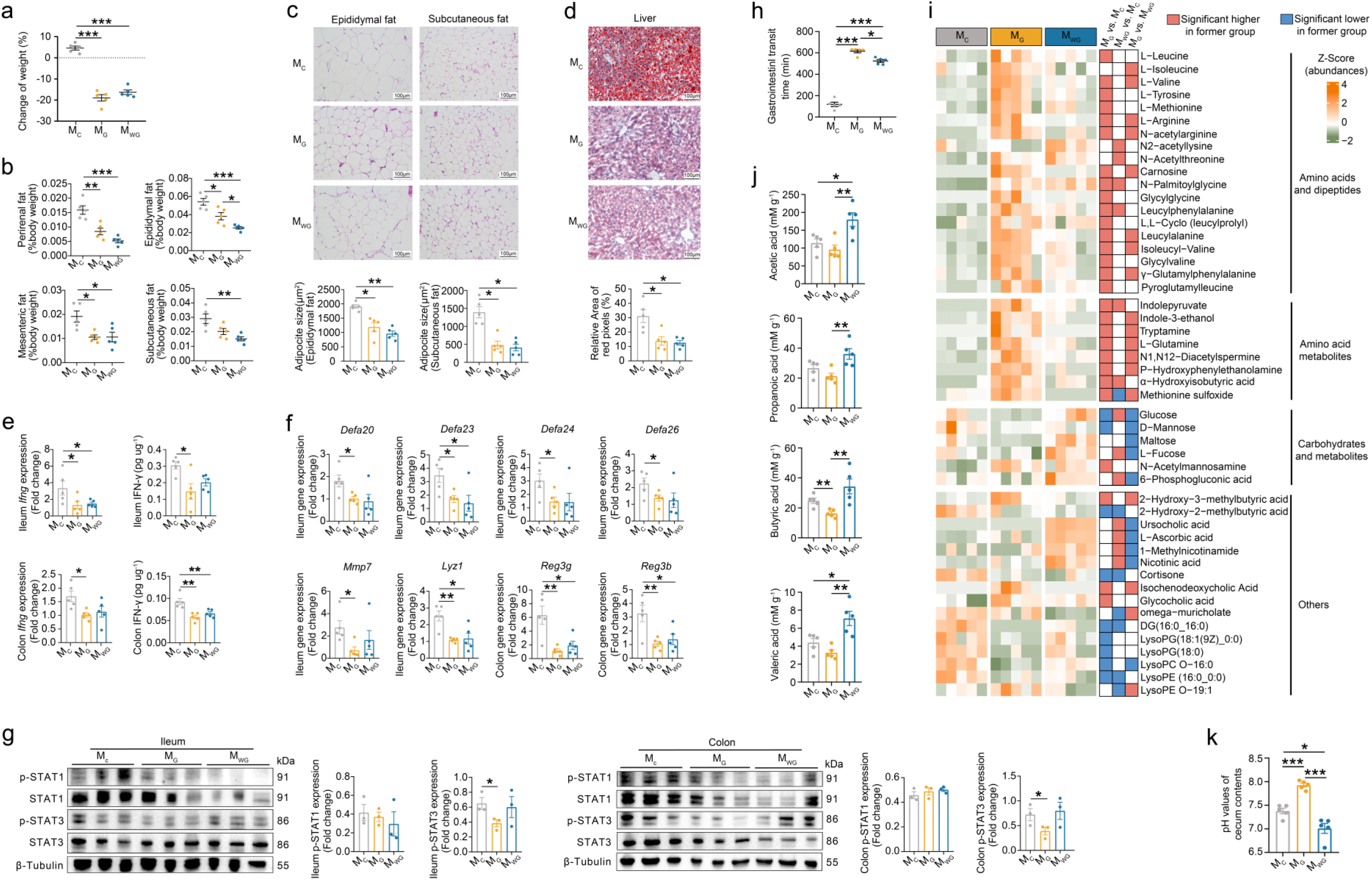
GLP-1 RA exacerbated gut microbiome dysbiosis by downregulating the expression of antimicrobial peptides and delaying gut transit time to alter the nutritional environment in the intestine. **a**, Changes in body weight from pre-intervention to post-intervention. **b**, Weight coefficients of adipose tissues. **c**, Representative H&E-stained sections of epididymal and subcutaneous fats (scale bar = 100 μm) and calculated mean adipocyte area. **d**, Representative Oil red o-stained liver sections (scale bar = 100 μm) and calculated relative area of red pixels. **e**, Expression of the IFN-γ gene and protein in the intestine measured by RT ‒ qPCR and ELISA, respectively. **f**, Relative expression of antimicrobial peptides genes in the intestine measured by RT‒qPCR. **g**, Representative western blot images and quantitative analysis of STAT1/3 phosphorylation in the intestine (*n*=3 per group). **h**, Gastrointestinal transit time among groups. **i**, Heatmap shows the abundances (Z scored) of differentially abundant colon content metabolites among groups. **j**, Concentrations of short-chain fatty acids in cecal contents among groups. **k**, pH values of cecum contents among groups. In all panels except **g**, *n*=5 per group. In all panels except **i**, data are presented as the mean ± s.e.m. In all panels, statistical analyses were performed using one-way ANOVA with Tukey’s post hoc test (**a**, **b**, **e**, **f**, **g**, **i**) or Kruskal‒Wallis test with Dunn’s post hoc test (**c**, **d**, **h**, **j**, **k**). \**P* < 0.05, \*\**P* < 0.01, \*\*\**P* < 0.001. M_C_, control group. M_G_, semaglutide intervention group. M_WG_, dietary fibre comminated with semaglutide intervention group.

A previous study demonstrated that GLP-1 RA targets its receptor of gut IELs and modulates the cAMP/PKA pathway to reduce IFN-γ production^21^. In DIO mice after 4 weeks of semaglutide treatment, we found that the levels of IFN-γ in the ileum and colon were significantly lower in the M_G_ group than in the M_C_ group (Figure 5e). Within the intestinal tract, the host surveils the gut microbiota via the production of diverse AMPs, while IFN-γ modulates AMPs generation^23^. We tested the expression of AMPs, including α-defensins, β-defensins, C-type lectin family, lysozyme and secretory phospholipase A_2_ in the ileum and colon of mice. Compared to the M_C_ group, the M_G_ group showed a significant decrease in the gene expression of *Defa20, 23, 24* and *26* (α-defensins), *Mmp7* (matrix metallopeptidase 7; responsible for activation of pro-α-defensins) and *Lyz1* (lysozyme) in the ileum and had lower levels of *Reg3γ* and *Reg3β* (C-type lectin family) in the colon (Figure 5f; Figure S8). Moreover, IFN-γ controls AMP-related genes expression by regulating the phosphorylation of the cytokine-responsive transcriptional activators STAT1/3. By western blotting, we found a significant decrease in the phosphorylation of STAT3 in the M_G_ group. (Figure 5g). These results suggest that semaglutide treatment induces weakened host immune surveillance over the gut microbiome in DIO mice, which already exhibit a dysbiotic state characterized by an elevated pathobiont ratio and proinflammatory signatures prior to drug administration.

Long gastrointestinal transit time (GTT), which represents a key therapeutic action of GLP-1 RAs, may be associated with alterations in gut microbiota metabolic profiles. We quantified GTT in DIO mice via oral gavage administration of methylcellulose-carmine red suspension and found that GTT was significantly longer in the M_G_ group than in the M_C_ group (Figure 5h). We performed LC‒MS-based untargeted metabolomic analysis with colonic contents and GC-based tests of SCFAs with cecal contents in DIO mice after 4 weeks of semaglutide treatment. Compared to the M_C_ group, mice in the M_G_ group had significantly higher levels of amino acids and dipeptides and lower levels of monosaccharides, reflecting an alteration in the availability of nutritional substrates for gut microbiota (Figure 5i). Moreover, mice in the M_G_ group had increased production from amino acid metabolism, including indole derivatives (e.g., indole-3-ethanol), amine compounds (e.g., P-hydroxyphenylethanolamine) and sulfur-containing metabolites (e.g., methionine sulfoxide), indicating proteolytic activity of the gut microbiome (Figure 5i; Table S10). On the other hand, the decrease in 6-phosphogluconic acid and SCFAs in the M_G_ group reflected reduced activity of the bacterial glycolytic pathway for carbohydrates (Figure 5i and 5j). These results indicate that GLP-1 RA treatment induces a shift in microbial metabolism from saccharolysis towards proteolysis. Notably, we also found that the pH value in the cecal contents of the M_G_ group was significantly higher than that of the M_C_ group (Figure 5k), which provides an environment that is more conducive to the proliferation of opportunistic pathogens.

Next, to test whether an increase in the intestinal properties of carbohydrates can mitigate adverse effects on the gut microbiota induced by semaglutide, we treated DIO mice with oral gavage of dietary fibre (same as that used in the clinical study; 500 mg/kg bodyweight/day) comminated with semaglutide injection (M_WG_ group). After 4 weeks of treatment, there was no significant difference between the M_WG_ and M_G_ groups in body weight change, weight of the fat pad, adipocyte size of adipose tissue, and lipide content of the liver (Figure 5a-5d). Moreover, M_WG_ mice exhibited reduced IFN-γ levels with concomitant downregulation of lysozyme and the C-type lectin family compared to the M_C_ group, which was similar to M_G_ mice (Figure 5e-5g; Figure S8). The GTT in the M_WG_ group was still much longer than that in the M_C_ group, although it was shorter than that in the M_G_ group (Figure 5h). These results indicate that dietary fibre intake does not compromise semaglutide-mediated regulation of body weight, IFN-γ/AMPs expression, and GTT in mammalian hosts. However, mice in the M_WG_ group had lower levels of amino acids, dipeptides and products from amino acid metabolism in the gut but higher levels of 6-phosphogluconic acid and SCFAs than those in the M_G_ group. Moreover, these amino acid-related metabolites showed no significant difference between the M_WG_ and the M_C_ group, while carbohydrates-related metabolites were much higher in the M_WG_ than in the M_C_ group (Figure 5i and 5j; Table S10). This reflected that fibre intake significantly enhanced saccharolysis and reduced proteolysis in the gut microbiota. Notably, we also found that the pH value in the cecal contents was significantly decreased in the M_WG_ group compared with the M_G_ and M_C_ groups (Figure 5k). On the other hand, the total bacterial load showed a lower trend in the M_WG_ than in the M_G_ group, and the whole structure of the gut microbiota was significantly different between the M_WG_ and M_G_ groups (Figure S7a and S7c). The opportunistic pathogens enriched by semaglutide treatment, such as ASVs belonging to *Escherichia−Shigella, Enterobacter* and *Enterococcus*, were significantly decreased in the M_WG_ group. Beneficial bacteria, such as ASVs from *Bifidobacterium pseudolongum* and *Lactobacillus johnsonii*, were significantly increased in the M_WG_ group compared with either the M_G_ or M_C_ group (Figure S7d; Table S9), suggesting that fibre induces alleviation of gut microbiota in DIO mice.

Taken together, GLP-1 RA-induced slow gut transit limited carbohydrate availability in the colon, leading microbial metabolism from saccharolysis towards proteolysis. Together with a decrease in AMPs-induced weakened host immune surveillance, GLP-1 RA thus exacerbates the dysbiosis of gut microbial composition and metabolism in DIO mice.

### GLP-1 RA in combination with dietary fibre attenuated weight regain following treatment cessation

To test whether mitigating the adverse effects of GLP-1 RA on the gut microbiota can reduce posttreatment weight rebound, we performed a 3-week follow-up experiment in DIO mice after treatment cessation of semaglutide with or without dietary fibre. After 3 weeks of treatment cessation, the regain of body weight, weight of the fat pad, adipocyte size of adipose tissue, lipid content of the liver, and levels of triglycerides in the liver and adipose tissue were significantly lower in M_WG_ mice than in M_G_ mice (Figure 6a-6e). This suggests that the intake of dietary fibre during semaglutide treatment can efficiently reduce post-treatment weight rebound.

**Figure 6.**
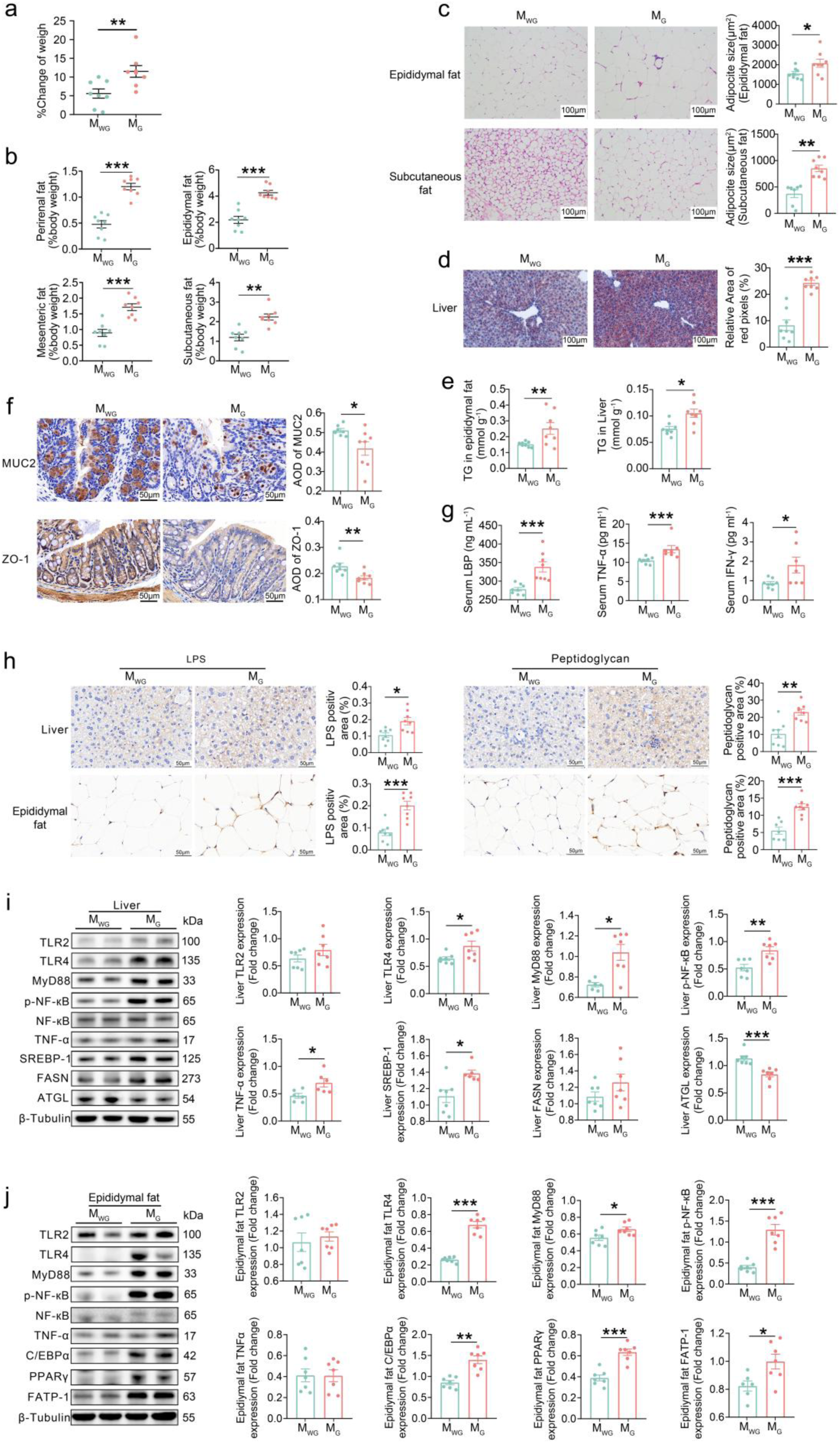
GLP-1 RA in combination with dietary fibre attenuated weight rebound posttreatment cessation. **a**, Changes in body weight from treatment cessation to the 3^rd^ week post-cessation. **b**, Weight coefficients of adipose tissues. **c**, Representative H&E-stained sections of epididymal and subcutaneous fats (scale bar = 100 μm) and calculated mean area of adipocytes. **d**, Representative Oil red o-stained liver sections (scale bar = 100 μm) and calculated relative area of red pixels. **e**, Triglyceride (TG) contents of liver and epididymal fat. **f**, Representative images of MUC2 and ZO-1 immunohistochemical staining in the colon (scale bar = 50 μm) and calculated average optical density (AOD). **g**, Serum levels of lipopolysaccharide binding protein (LBP), TNF-α and IFN-γ measured by ELISA. **h**, Representative images of lipopolysaccharide (LPS) and peptidoglycan immunohistochemical staining in liver and epididymal fat (scale bar = 50 μm) and calculation of the positive areas. **i** and **j**, Representative western blot images and quantitative analysis of key regulators in the TLRs/MyD88/NF-κB inflammatory pathway and lipid metabolism in liver (**i**) and epididymal fat (**j**). *n*=7 per group. In **a** to **h**, *n*=8 per group. In all panels, data are presented as the mean ± s.e.m., and statistical analyses were performed using unpaired two-tailed Student’s *t* test (**a**, **b**, **i**, **j**) or two-tailed Mann‒Whitney U test (**c**, **d**, **e**, **f**, **g**, **h**). \**P* < 0.05, \*\**P* < 0.01, \*\*\**P* < 0.001. M_G_, semaglutide intervention group. M_WG_, dietary fibre comminated with semaglutide intervention group.

In parallel, we found that the structure of the gut microbiota still showed differences between the M_WG_ and M_G_ groups even 3 weeks after treatment cessation, and dietary fibre-induced enrichment of beneficial bacteria (e.g., *Bifidobacterium pseudolongum*) and a decrease in opportunistic pathogens (e.g., *Enterobacter* sp.) were still preserved in the M_WG_ group (Figure S9; Table S11). Consequently, compared to the M_G_ group, the M_WG_ group had higher levels of ZO-1 and MUC2 in the colon (Figure 6f). Moreover, LBP and proinflammatory cytokines, such as TNF-α and IFN-γ, were significantly lower in M_WG_ mice (Figure 6g). These results indicate that the mice that received semaglutide in combination with dietary fibre had better gut barrier function and lower systemic inflammation after treatment cessation than their companions treated with semaglutide alone.

Next, we also found significantly lower accumulation of LPS and peptidoglycan (Figure 6h), downregulation of TLR4 and MyD88, decrease of nuclear translocation of NF-κB, and reduction of TNF-α in liver and adipose of M_WG_ mice than M_G_ mice (Figure 6i and 6j). Subsequently, compared to the M_G_ group, the NF-κB/SREBP-1/FASN-related lipogenesis pathway was downregulated, and the ATGL-related lipolysis pathway was upregulated in the livers of M_WG_ mice, while the C/EBPα/PPARγ/FATP-1-related uptake of long-chain fatty acids pathway was decreased in adipose tissue (Figure 6i and 6j).

These results indicate that mitigating the adverse effects of GLP-1 RA on gut microbiota by dietary fibre intake can efficaciously reduce posttreatment weight regain, which reinforces the pivotal role of dysbiotic gut microbiota in GLP-1 RA cessation-associated weight rebound and offers a clinically actionable solution.

## Discussion

GLP-1 RAs are increasingly utilized for the treatment of obesity. In the current study, we demonstrate that by modulating gut AMPs expression to weaken host immune surveillance over the gut microbiota and delaying GTT to change microbiota-available nutritional substrates, semaglutide amplified the proinflammatory signature of the gut microbiota in obese individuals, which contributed to lipid accumulation in the liver and adipose tissue following treatment cessation. Moreover, dietary fibre co-administration with semaglutide alleviated proinflammatory microbiota to reduce posttreatment weight regain.

Clinically significant short-term weight reduction can be achieved through intensive lifestyle modification, pharmacotherapy, or bariatric surgery, but weight regain following medication discontinuation poses a significant concern^12,24,25^. This ‘yo-yo’ effect may not only be attributable to an obesogenic metabolic memory of adipose tissue that persists but also depend on whether obesity-associated gut microbial dysbiosis has been therapeutically corrected^15,26^. In the current clinical trial and mouse experiment, we showed that posttreatment weight regains associated with semaglutide aggravated gut microbial dysbiosis, which can induce gut barrier dysfunction, chronic and systemic inflammation and adiposity expansion. Our data indicate that GLP-1 RA conferring host adiposity reduction or metabolic improvement does not necessarily translate to amelioration of gut microbial dysbiosis. Lifestyle modifications, particularly dietary interventions such as the high-fibre diet employed in this study, not only improve metabolism in patients with obesity but also ameliorate gut microbiota dysbiosis. This underlying mechanistic benefit provides a plausible explanation for the findings of a recent meta-analysis, which revealed that following intervention cessation, weight regain among subjects receiving lifestyle modifications was significantly lower than in those treated with GLP-1 RAs, independent of initial weight loss^13^. This underscores the necessity of deliberate microbial consideration when implementing non-microbiota-targeted therapeutic modalities for diseases management, given their potential to inadvertently reshape the gut ecosystem. Therapeutically remodelled gut microbiota exerts sustained effects on the host even after discontinuation of the primary intervention, persisting beyond the cessation of energy deficit induction or pharmacodynamic actions on metabolic pathways.

Wong *et al*. demonstrated that GLP-1 RA reduced IFN-γ production from gut IELs through modulation of the cAMP/PKA pathway, which is not essential for metabolic homeostasis^21^. They showed that the GLP-1 receptor of gut IEL controls a subset of GLP-1 RA actions^21^. Our current work suggests that the decrease in IFN-γ induced a reduction in AMPs in the gut, suggesting that weakened host immune surveillance over the gut microbiome is one of the critical impacts of GLP-1 RAs in the mammalian intestine. It must be appreciated that the gut microbiota in obese individuals already exhibits distinct structural and functional profiles from healthy counterparts, particularly characterized by enhanced endotoxin-generating pathobionts and reduced beneficial metabolite producers^17,27^. A decrease in intestinal AMPs may provide a condition for further overgrowth of opportunistic pathogens^28^. In the context of GLP-1 RA interventions, the beneficial effects of this drug selective restraint of not only local and systemic T-cell-induced inflammation but also LPS-induced inflammation^9,21^, may be sufficient to offset the adverse impacts of the proinflammatory gut microbiome shaped by GLP-1 RA on the host. However, this proinflammatory gut microbiome persists in the host following treatment cessation, contributing to sustained systemic chronic inflammation post-discontinuation.

Delaying gastrointestinal transit induced by GLP-1 RAs is a pivotal physiological effect related to a reduction in body weight^29^. However, evidence from diverse disease studies has shown that a longer gut transit time is associated with a higher fecal pH, reduced fecal water content, higher microbial cell density and diversity, and a shift in microbial metabolism from saccharolysis towards proteolysis^30^. Slow gut transit limits carbohydrate availability in the colon, favouring bacteria that can use other sources of energy, such as dietary or host-derived proteins^31^. During gut microbial community assembly, dietary electron donors govern the availability of ecological positions within the microbiota^32^. Moreover, the nature of the microbial products also changes the physicochemical properties of the colonic environment, for example, by changing pH and thus altering the microbial composition and metabolism^33^. While saccharolysis by the gut microbiota gives rise to SCFAs that are beneficial for the host and a source of energy for the colonocytes^34^, proteolysis can lead to the accumulation of compounds such as branched-chain fatty acids, phenols, indoles and ammonium that are generally considered detrimental for health^35^. Given the host’s lack of enzymes to metabolize dietary fibre^36^, the undigested fibre reaches the colon, thereby correcting the unfavourable colonic nutritional environment induced by longer gut transit time. This increase in available carbohydrate substrates for bacterial fermentation leads to an improvement in the gut microbiota. Consequently, this mitigates the risk of weight regain upon discontinuation of GLP-1 receptor agonist therapy.

Our study suggests that with the development of novel anti-obesity pharmacotherapies, we should not only focus on evaluating their efficacy and elucidating their mechanisms of action in the human body but also give due attention to their effects on the gut microbiome. The changes in the gut microbiome that occur during pharmacological treatment may have intimate connections with treatment responsiveness, adverse event profiles, and the risk of weight regain upon treatment discontinuation. Alongside the use of these anti-obesity medications, the adoption of dietary and lifestyle interventions aimed at improving the gut microbiome, such as the incorporation of dietary fibre, may hold the potential to achieve more effective and durable weight loss benefits. By concurrently addressing both the pharmacological and microbiome-modulating aspects of obesity management, we may be able to optimize the long-term outcomes for patients undergoing anti-obesity treatment.

### Limitations of the study

One limitation of this study is that we lacked serum and fecal samples at the 14^th^ week post-cessation. We could not further confirm in a human cohort that the proinflammatory microbiota shaped by GLP1-RA persisted in the host and promoted weight regain by inducing chronic low-grade systemic inflammation in a clinical trial. Additionally, future studies are still needed to conduct interventions combining GLP-1 RA with dietary fibre in human cohorts, aiming to verify that dietary fibre reduces posttreatment weight regain by mitigating the adverse effects of GLP-1 RA on the gut microbiota.

## Resource availability

### Lead contact

Further information and requests for resources and reagents should be directed to and will be fulfilled by the lead contact, Chenhong Zhang; Xiaoying Ding

### Materials availability

All unique reagents generated in this study are available from the lead contact upon request.

### Data and code availability

To protect participant privacy, access to and use of deidentified individual-level clinical data should be requested from the corresponding authors. Generally, all requests for access to data will be responded to within 1 month. The clinical data can only be used for non-commercial scientific research purposes. The raw sequence data have been deposited in the Genome Sequence Archive (GSA) database in the National Genomics Data Center (NGDC) (https://ngdc.cncb.ac.cn/gsa), and the accession numbers of the clinical study, the FMT animal trial, and the semaglutide and dietary fibre interventions animal trial are CRA033354, CRA033365, and CRA033219, respectively. Source Data are provided with this paper. No custom code was used in this paper. All publicly available codes and tools used to analyse the data are reported and referenced in the Methods.

## Acknowledgements

The authors sincerely thank the medical staff and study participants who took part in the trial. This work was supported by the National Key Research and Development Project (No. 2022YFF1100103), the National Natural Science Foundation of China (81870594) and the Clinical Research Plan of Shanghai Hospital Development Center (No. SHDC2020CR1016B). This work was also supported by Adfontes (Ningbo) Co., Ltd. The sponsor was not in the study design, data collection, analysis, writing of the manuscript, or the decision to submit the article for publication.

## Author contributions

C.Z., and X.D. conceived and designed the clinical study. X.D., Y.L., Y.M., Y.S., B.P., F.Z. and Y.P. made clinical diagnoses, recruited participants and performed interventions. X.D., Y.L., Y.M., Y.S., B.P., F.Z., R.Z., Y.Z., and H.M. collected clinical samples and questionnaires and conducted clinical tests. R.Z. and Y.Z. provided dietary counselling and prepared and delivered a high-fibre diet to the subjects. D.S. and C.Z. analysed clinical data and metagenome data. C.Z. and D.S. conceived and designed the animal study. D.S. conducted animal feeding and intervention. D.S. and Y.H. carried out short-chain fatty acid profiling and data analysis. D.S., and Z.W. performed histopathological analysis. D.S., Z.W., and Z.F. extracted fecal DNA and tissue RNA from the mice. D.S. carried out animal microbial and metabolome data analysis, RT‒qPCR, and western blotting. C.Z. and D.S. wrote the manuscript.

## Declaration of interests

Chenhong Zhang is the co-founder of Adfontes (Ningbo) Co., Ltd. The remaining authors declare no conflict of interest.

### **STAR**★**METHODS**

### Key resources table

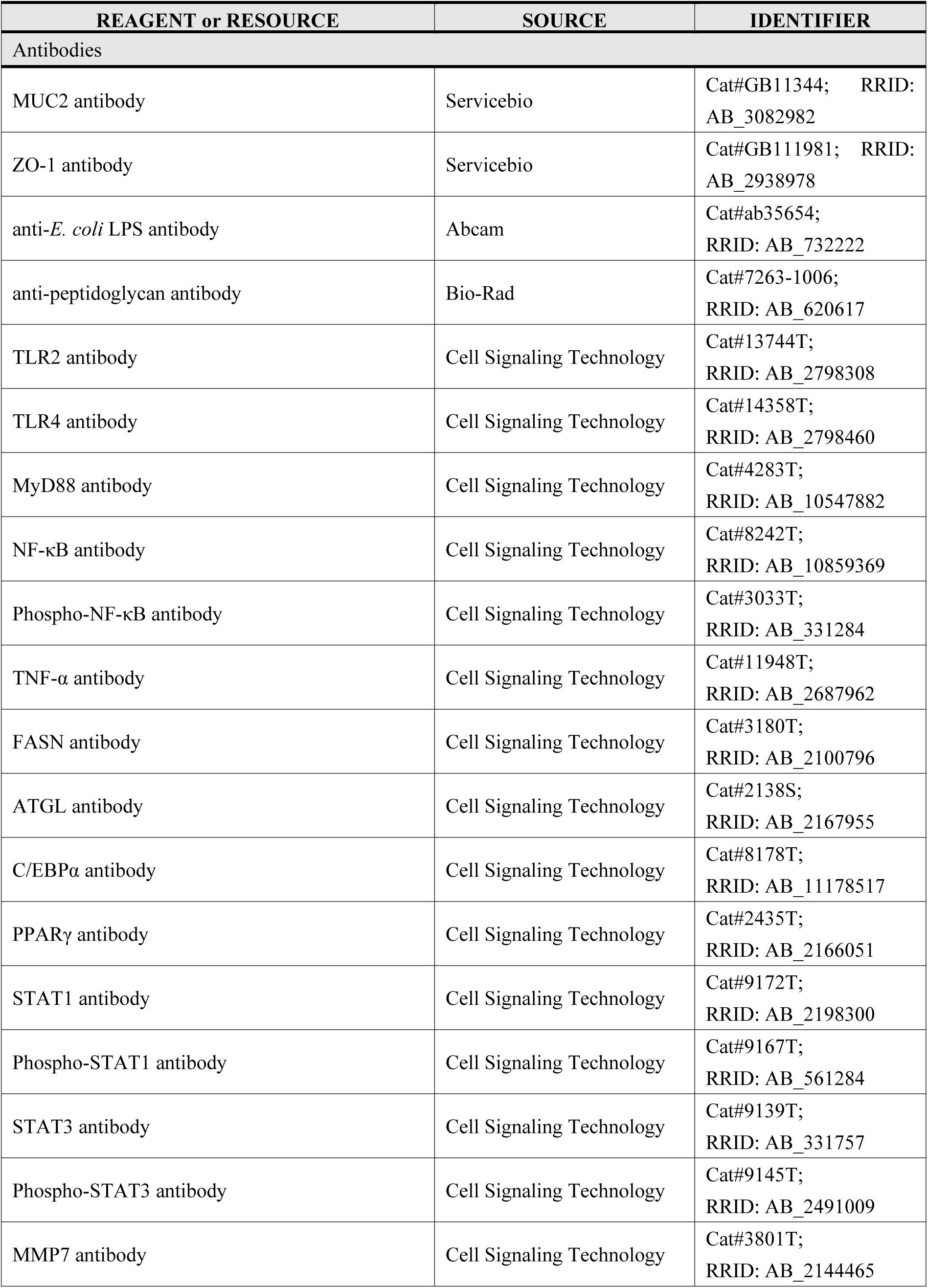
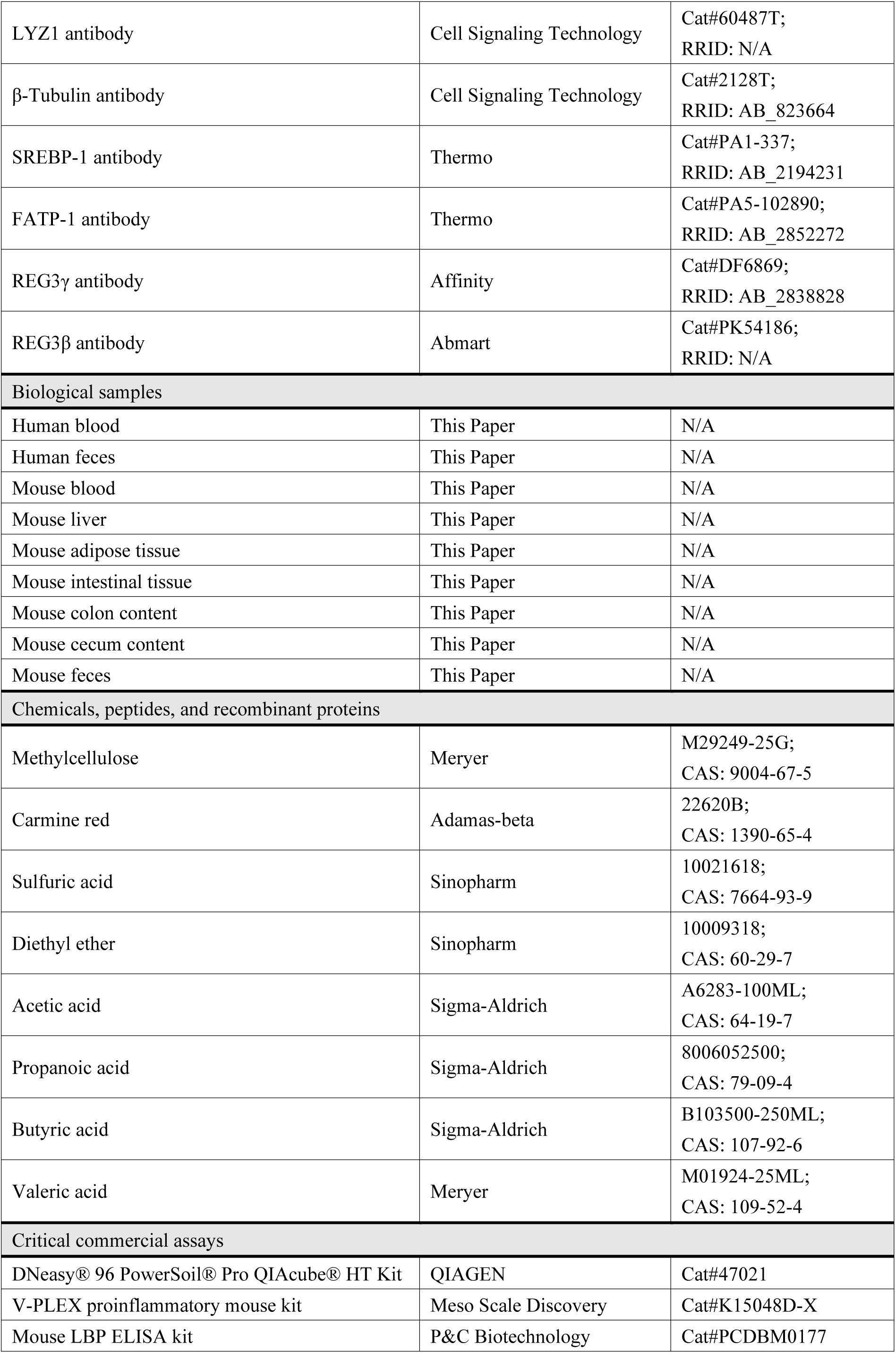
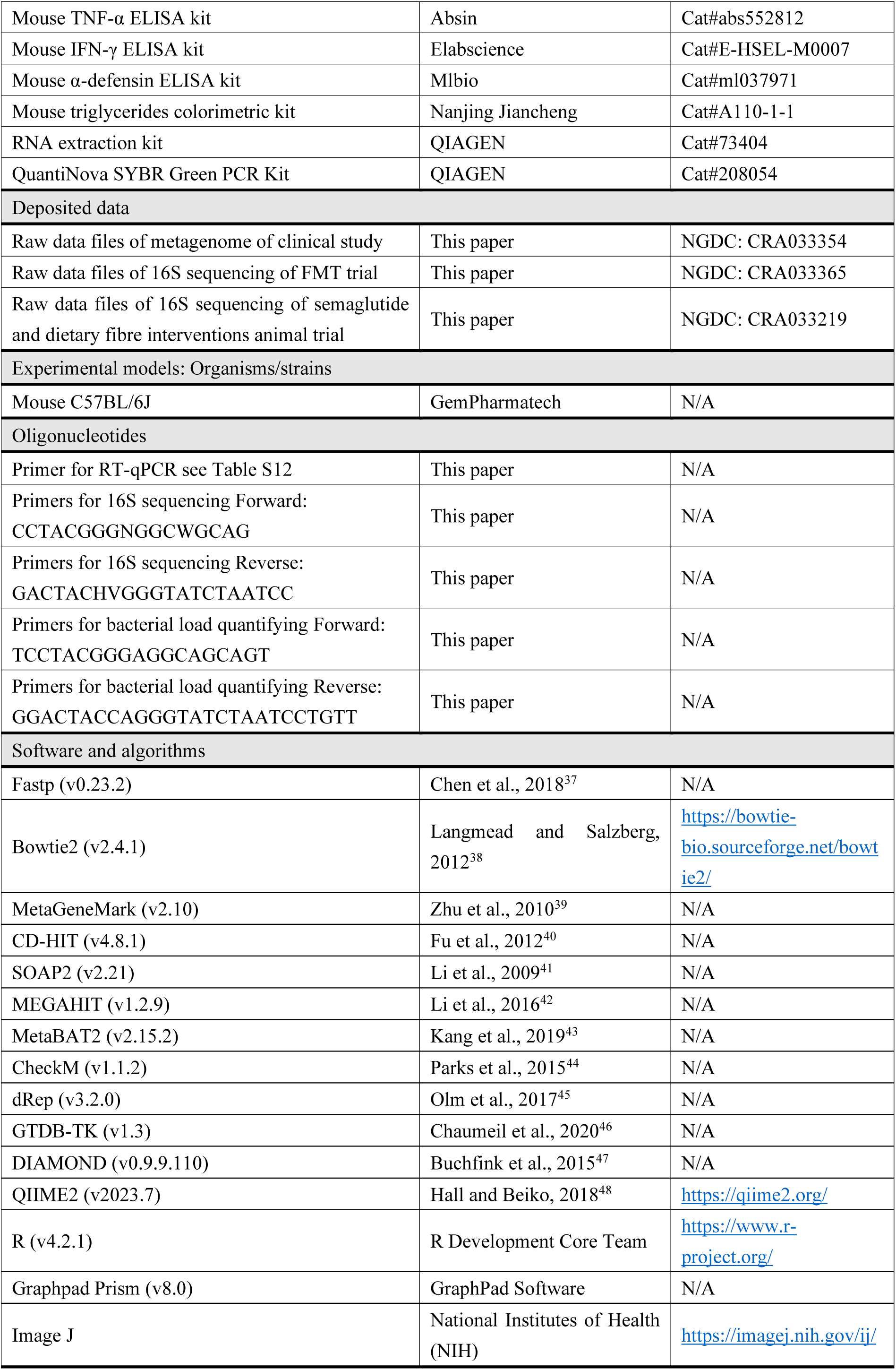

### Experimental model and study participant details

#### Clinical Study

The semaglutide or high-fibre diet intervention study is an open-label, multicenter clinical trial for overweight and obese subjects. Its protocol was approved by the Shanghai General Hospital Ethics Committee (ID: [2022]188), conducted with the Declaration of Helsinki. All subjects provided written informed consent. The trial was registered in the Chinese Clinical Trial Registry: ChiCTR2200066014 (https://www.chictr.org.cn/).

##### Study Design

The study design and subject flow are shown in Figure 1a. The semaglutide and high-fibre diet intervention lasted 12 weeks, followed by a 26-week body weight change follow-up after intervention cessation.

###### Outcomes

The primary outcome was the percentage change in weight after the intervention. Secondary outcomes were the percentage changes in BMI, waist and hip circumferences, waist-to-hip ratio, blood pressures, body fat and lean mass, glucose, HbA1c, insulin, c-peptide, HOMA2-IR, lipid, liver and kidney function parameters, anxiety and depression scores. Exploratory outcomes were the change in gut microbiota after intervention and the weight change after intervention cessation.

###### Sample size estimation

Sample size was calculated using the standard formula for non-inferiority trials. The weight changes at 9 weeks showed a mean reduction of -6.0%±4.0% in the dietary fibre intervention group^49^. The weight changes at 12 weeks showed a mean reduction of -6.6%±4.0% in the semaglutide intervention group^11,50^. Sample size calculation (α=0.05, power=90%, 1:1 allocation, 10% dropout rate, non-inferiority margin = 3.3%) yielded 34 per group. Therefore, we planned to enrol at least 34 subjects per group in this study. Given the poor adherence and high dropout rate of dietary intervention, the sample size for the high-fibre diet intervention group should be greater than that for the semaglutide intervention group.

###### Inclusion and exclusion criteria

Subjects were recruited between December 3^rd^ 2022, and December 16^th^ 2022, at Shanghai General Hospital (Shanghai, China), Qidong People’s Hospital (Qidong, China), and Affiliated Hospital of Jiangnan University (Wuxi, China).

The inclusion criteria were age between 18 and 60 years and BMI ≥ 24 kg/m^2^. The exclusion criteria were pregnancy or lactation; diagnosed with diabetes, hepatic, kidney and gastrointestinal diseases; allergy to intervention drug or food; weight loss through medication or surgery within 3 months pre-enrollment; and used antibiotics within 2 weeks pre-enrollment.

###### Randomization

A completely randomized design was employed to assign subjects into semaglutide or high-fibre diet intervention groups in a 1:1.5 ratio using the R package randomizr (v1.0.0)^51^.

###### Intervention

Subjects received either once-weekly subcutaneous injection semaglutide (Ozempic, Novo Nordisk, Denmark) or the WTP diet (45 g dietary fibre/day/person^34^ as staple replacement) intervention for 12 weeks. The semaglutide dose was as follows: 0.25 mg (weeks 1-2), 0.5 mg (weeks 3-4), and 1.0 mg (weeks 5 onwards). The WTP diet was based on wholegrains, traditional Chinese medicinal foods and prebiotics, containing wheat bran, oat bran, *Coix lachrymal-jobi* L., *Fagopyrum tataricum* L., *Hordeum vulgare var. coeleste* L., *Salvia Hispanica* L., *Chenopodium quinoa* Willd., *Secale cereale* L., *Setaria italica* L., resistant starch, inulin, fructo-oligosaccharides and xylo-oligosaccharides. The nutritional composition of the WTP diet is detailed in Table S1.

###### Samples collection and bio-clinical parameters measurement

At baseline and post-intervention, the following samples and bio-clinical parameters were collected and measured: anthropometric parameters; fasting blood sampling for glucose, HbA1c, insulin, C-peptide, lipids, and liver/kidney function parameters; a 75 g oral glucose tolerance test (120-min blood sampling for postprandial glucose, insulin, and C-peptide); fecal sample for gut microbiota analysis; and questionnaires (food frequency questionnaires, self-rating depression scale, and self-rating anxiety scale). Weight was measured at the 26-week follow-up. Details are provided in the Supplementary Methods.

#### Animal Study

##### FMT animal trial

The trial was conducted at the laboratory animal center of GemPharmatech Co., Ltd. (Nanjing, China), and the procedures were approved by the Institutional Animal Care and Use Committee (IACUC) of GemPharmatech Co., Ltd. (GPTAP20231122-4, GPTAP20240520-5). Six-week-old germ-free male C57BL/6J mice received oral gavage with a 200-µl fecal slurry (3 times/week) for 2 weeks. Mice were randomly divided into M_FW_ and M_FG_ groups and inoculated with pos-tintervention fecal slurry from two donors of the W and G groups, respectively. All mice were fed a normal chow diet (AIN-93G, SYSE Bio-tech. Co., Ltd., China) and were housed in plastic isolators maintained at 20-26°C with 40–70% humidity and a 12-hour light/dark phase cycle. The experiments were conducted in two independent trials: 5 mice per group in trial 1 and 10 mice per group in trial 2, and the batch effect was adjusted using the R package sva (v3.44.0)^52^.

##### Semaglutide and dietary fibre interventions animal trial

The trial was conducted at the laboratory animal center of Shanghai Jiao Tong University (Shanghai, China), and the procedures were approved by the IACUC of Shanghai Jiao Tong University (A2025093). Mice were housed in a specific pathogen-free facility maintained at 22.0 ± 1°C with 30–70% humidity and a 12-hour light/dark phase cycle. Eight-week-old SPF male C57BL/6J mice were fed a Western-style diet (D12079B, SYSE Bio-tech. Co., Ltd., China) for 4 weeks and then received 4 weeks of interventions: subcutaneous injection of semaglutide (0.1 mg/kg body weight/every three days, M_G_ group); semaglutide plus oral gavage of dietary fibre (same as that used in the clinical study; 500 mg/kg bodyweight/day^53^, M_WG_ group); and normal saline injection plus sterile water gavage as a control (M_C_ group). The mouse semaglutide dose was equivalent to 1.0 mg/kg in adults via body surface area-based dose translation. Post-intervention, the mice in the M_WG_ and M_G_ groups were fed a normal chow diet (AIN-93G, SYSE Bio-tech. Co., Ltd., China) for 3 weeks.

### Method details

#### Animal experiments

##### Histopathological analysis

Liver, epididymal fat and subcutaneous fat tissues were fixed with 4% paraformaldehyde, embedded in paraffin, sectioned (4-μm thickness), and stained with hematoxylin and eosin (G1003, Servicebio, China). The liver was stained with Oil Red O (O0625, Sigma‒Aldrich Ltd., USA). Digital images of sections were acquired with a Leica DMRBE microscope. The adipocyte mean area and liver lipid droplet relative area were measured using ImageJ software. The nonalcoholic fatty liver disease (NAFLD) score was determined by two blinded pathologists according to the method described by Kleiner DE *et al*^54^.

##### Immunohistochemical staining

Colon sections were incubated with MUC2 antibody (1:500 dilution) and ZO-1 antibody (1:200 dilution). The liver and epididymal fat sections were incubated with anti-*E. coli* LPS antibody (1:50 dilution) and anti-peptidoglycan antibody (1:50 dilution). Stained sections were captured with an Eclipse E100 microscope (Nikon, Japan), and at least three fields per slide were randomly selected for calculating average optical density (AOD) values of MUC2 and ZO-1 and positive areas (%) of LPS and peptidoglycan by ImageJ software.

##### Biochemical and immunological assays

For the FMT animal trial, serum proinflammatory cytokines (including IL-1β, IL-2, IL-4, IL-5, IL-6, IL-10, IL-12, TNF-α, IFN-γ, and KC) were measured by the meso scale discovery-electrochemiluminescence method following the instructions of the V-PLEX proinflammatory mouse kit using the MESO QuickPlex SQ 120MM instrument (Merck Sharp & Dohme, USA). Serum LBP levels were measured by enzyme-linked immunosorbent assays (ELISA). Triglyceride levels in liver and adipose tissues were measured by the glycerophosphate oxidase-peroxidase-aminophenazone-phenol (GPO-PAP) method. For the semaglutide and dietary fibre interventions animal trial, serum levels of LBP, TNF-α, and IFN-γ and intestinal levels of IFN-γ and α-defensin were measured by ELISA. Triglyceride levels in liver and adipose tissues were measured by the GPO-PAP method. The kits used were shown in key resources table.

##### Quantitative PCR with reverse transcription (RT–qPCR)

The total RNA of tissue samples was extracted using an RNA extraction kit (73404, QIAGEN, Germany) and reverse transcribed into cDNA using SuperScript-III reverse transcriptase (18080051, Thermo Fisher, USA). RT–qPCR was performed using the QuantiNova SYBR Green PCR Kit on a qTOWER 3G Real-time PCR system (Analytik Jena, Germany). Gene expression levels were determined using the comparative CT method (2^-ΔΔ*Ct*^), with *Actb* serving as the reference gene for liver and colon and *Rplp0* serving as the reference gene for adipose tissue. All primer sequences are listed in Table S12.

##### Western blotting

The proteins of liver and epididymal fat samples were extracted using RIPA buffer (P0013B, Beyotime, China) containing protease and phosphatase inhibitors (P1045, Beyotime, China). Protein samples were resolved by SDS–PAGE (P0015B, Beyotime, China) and immunoblotted onto polyvinylidene difluoride membranes. Immunoreactivity was detected using enhanced chemiluminescent autoradiography (180-501, Tanon, China). Band quantification was performed using ImageJ software. The antibodies were shown in key resources table. All antibodies were diluted 1:1000.

##### Gastrointestinal transit time (GTT) measurement

Mice received 0.3 ml of 0.5% methylcellulose solution containing 6% carmine red orally. Then, the mice were allowed food and water freely until the first red fecal pellet appeared. The GTT was defined as the time from oral administration to the appearance of the first red fecal pellet.

##### Short-chain fatty acids (SCFAs) measurement

The SCFA concentrations of cecum contents were determined using gas chromatography spectrometry (GC) as previously described^34^. Briefly, 100 mg cecum contents were homogenized with 1 mL normal saline (pH 7.0) and centrifuged, and the supernatant was collected and filtered for sterilization. The pH value of the supernatant was measured using an FE28-pH meter (Mettler Toledo, Switzerland). Sulfuric acid (50%) was added to the supernatant for acidification treatment, and the organic acids were extracted using diethyl ether and then measured on a Thermo TSQ8000 (Thermo Fisher, USA).

##### Untargeted metabolomics analysis

Serum and cecal contents from the FMT trial (trial 1) were sent to Shanghai Applied Protein Technology Co., Ltd. (Shanghai, China), and the colon contents from the semaglutide and dietary fibre intervention trial were sent to Panomix Biomedical Tech Co., Ltd. (Suzhou, China) for untargeted metabolome profiling. Liquid chromatographic separation was performed on a Vanquish UHPLC System (Thermo Fisher, USA). Mass spectrometric detection of metabolites was carried out using a Q Exactive (Thermo Fisher, USA) equipped with an ESI ion source. Details are provided in the Supplementary Methods.

Orthogonal partial least squares discriminant analysis (OPLS-DA) and PLS-DA were performed using the R package ropls (v1.28.2)^55^. Differentially abundant metabolites were identified using thresholds of variable importance in projection (VIP) > 1, *P* < 0.05 (two-tailed Student’s *t* test or one-way ANOVA with Tukey’s post hoc test), and absolute value of Log2(fold change) > 1 (only for cecal content metabolites).

##### Quantification of total bacterial load

The total bacterial load was quantified by qPCR. Briefly, qPCR was performed with a QuantiNova SYBR Green PCR Kit on a qTOWER 3G Real-time PCR system (Analytik Jena, Germany). A standard curve was generated using serial dilutions of a plasmid containing the full-length 16S rRNA gene from *Blautia obeum*.

#### Gut microbiota analysis

##### Shotgun metagenomic sequencing and analyses of clinical study

###### Metagenomic sequencing and data quality control

DNA was extracted from fecal samples as previously described^56^ and sequenced using the Illumina NovaSeq 6000 platform (Illumina, USA). Libraries were constructed with an approximately 500 bp insert size, followed by high-throughput sequencing for 150 bp paired-end reads (forward and reverse). Fastp (v0.23.2)^37^ was used to preprocess the raw data and obtain clean data. Then, clean data were blasted to the human genome reference database GRCh38 using Bowtie2 (v2.4.1)^38^ to filter out the reads that originated from the host (--end-to-end, --moresensitive, -I 200, -X 500). Details are provided in the Supplementary Methods.

###### Non-redundant gene construction and abundance calculation

Open reading frame (ORF) prediction was performed with MetaGeneMark (v2.10)^39^. CD-HIT (v4.8.1)^40^ was used to remove redundant sequences to obtain a non-redundant initial gene set. Per-sample gene read counts were calculated by comparing clean data to the initial gene set using SOAP2 (v2.21)^41^. The final non-redundant gene set was obtained by filtering genes with < 2 reads per sample. Non-redundant genes abundances were calculated by comparing read counts to gene length.

###### De novo assembly, taxonomy annotation, and abundance calculation of genomes

De novo assembly was performed for each sample using MEGAHIT (v1.2.9; min contig is 500)^42^. The assembled contigs were further binned using MetaBAT2 (v2.15.2)^43^. High-quality metagenome-assembled genomes (HQMAGs) were defined as those with >90% completeness and <5% contamination as assessed by CheckM (v1.1.2)^44^. The assembled HQMAGs were further dereplicated by using dRep (v3.2.0)^45^. Taxonomic assignment of the genomes was performed using GTDB-TK (v1.3)^46^ with default parameters. Then, clean reads from each sample were mapped to the genome using Bowtie2 (v2.4.1)^38^. The relative abundances of HQMAGs were defined as the number of reads aligning to each genome normalized by genome size.

###### Functional annotation of HQMAGs and non-redundant genes

DIAMOND (v0.9.9.110)^47^ was used to compare the gene ORF protein sequences predicted by HQMAGs and non-redundant genes with the KEGG database (v2019.10), CARD database (v2023.10), CAZy database (v2018.01), and VFDB database (v2021.05) to obtain the functional information of HQMAGs and non-redundant genes. Details are provided in the Supplementary Methods.

##### 16S rRNA gene sequencing and analyses of animal study

DNA extraction from mice fecal samples were performed as previously described^57^. The DNA samples were sent to Shanghai Major Biomedical Technology Co., Ltd. (Shanghai, China) for 16S rRNA gene sequencing of the V3-V4 region. Shotgun sequencing was performed on an Illumina HiSeq platform (Illumina, USA). The QIIME2 pipeline (v2023.7)^48^ was employed to obtain amplicon sequence variants (ASVs). The taxonomy of ASVs was annotated using the SILVA database (v138). The relative abundances of ASVs were determined by downsizing the sequencing depth to a common level (20000 reads). The relative abundance of ASVs was then converted into absolute abundance based on the total bacterial load for animal gut microbiota analyses. Details are provided in the Supplementary Methods.

##### Gut microbiota diversity analyses

###### Microbiota diversity analyses in the clinical study

Alpha diversity was calculated by the R package vegan (v2.6-4)^58^. Adjusted principal coordinates analysis (aPCoA) and permutational multivariate analysis of variance (PERMANOVA) with 999 permutations based on Bray‒Curtis distance were performed using the R packages aPCoA (v1.3)^59^ and vegan (v2.6-4)^58^, respectively. Subject was regarded as a covariate for adjustment in aPCoA and PERMANOVA.

###### Microbiota diversity analyses in the animal study

Alpha diversity was calculated by QIIME2 (v2023.7)^48^. PCoA and PERMANOVA (999 permutations) based on Bray‒Curtis distance were performed using R packages stats (v4.2.1)^60^ and vegan (v2.6-4)^58^, respectively.

##### Differentially abundant HQMAGs or ASVs identification

The Boruta random forests algorithm (1000 permutations, Bonferroni-adjusted *P* values) was performed using the R package Boruta (v9.0.0)^61^ to identify the discriminative HQMAGs or ASVs between different groups or pro- and post-intervention within the same group. Then, two-tailed Mann‒Whitney U tests (Inter-group) or two-tailed Wilcoxon matched-pair signed-rank tests (Intra-group) were conducted on the above selected HQMAGs or ASVs to further screen out those with significant abundance differences.

### Quantification and statistical analysis

#### Statistical analysis of the clinical study

Comparisons of proportion differences of categorical variables between groups were performed using Pearson’s chi-squared test; inter-group comparisons at the same timepoint were performed using a two-tailed Mann‒Whitney U test; intra-group comparisons at different timepoints were performed using a two-tailed Wilcoxon matched-pair signed-rank test. At baseline, a generalized linear model was used to assess the relationships between body weight and other variables. For comparing group differences in variables significantly related to body weight, two-tailed ANCOVA was adjusted for baseline body weight. Statistical analysis and plots were performed using R (v4.2.1).

#### Statistical analysis of the animal study

The Shapiro–Wilk test and Kolmogorov–Smirnov test were used to assess the data distribution normality. For normally distributed data (*P* > 0.05), two-group comparisons used two-tailed unpaired Student’s *t* test; multigroup comparisons used one-way ANOVA with Tukey’s post hoc test. For nonnormally distributed data (*P* ≤ 0.05), two-group comparisons used the two-tailed Mann–Whitney U test; multigroup comparisons used the Kruskal‒Wallis test with Dunn’s post hoc test. Statistical analysis and plots were performed using GraphPad Prism (v8.0) and R (v4.2.1). ROUT (Q = 1%) was used to test each group of data and eliminate outliers.

## Supplementary Information

### 1. Supplementary Methods

### 2. Supplementary Figures 1 to 9

Figure S1 The differentially abundant HQMAGs and functional features of the microbiome between the G and W groups or pro- and post-intervention within the same group.

Figure S2 The gut microbiome of the recipient mice was similar to that of corresponding human donors.

Figure S3 Gut microbiota in GLP-1 RA recipients led to lipid accumulation in germ-free mice in wo independent trials.

Figure S4 The differences in cecal content and serum metabolomes between MFG and MFW groups. Figure S5 The gut microbiota in GLP-1 RA recipients induces gut barrier dysfunction, activates he TLRs/MyD88/NF-κB inflammatory pathway, drives lipogenesis and suppresses lipolysis in mice in two independent trials.

Figure S6 The gut microbiota in GLP-1 RA recipients induced systemic inflammation, driving ipogenesis and suppressing lipolysis.

Figure S7 GLP-1 RA attenuated systemic inflammation while exacerbating gut microbiota dysbiosis in obese mice, whereas dietary fibre improved dysbiosis.

Figure S8 GLP-1 RA downregulated the expression of antimicrobial peptides by reducing the expression of IFN-γ in the intestine.

Figure S9 The gut microbiota of the MG and MWG maintains end-of-intervention characteristics at the 3rd week post-cessation.

### 3. Supplementary Tables 1 to 12

Table S1 Nutritional composition of the WTP diet.

Table S2 The baseline characteristics of the clinical study.

Table S3 The characteristics of dietary factors at baseline and after intervention. Table S4 Primary and secondary outcomes.

Table S5 Clinical parameters after intervention.

Table S6 The differentially abundant HQMAGs within or between the G and W groups in the clinical study.

Table S7 The differentially abundant ASVs between M_FG_ and M_FW_ groups.

Table S8 The differentially abundant metabolites of cecal content and serum between M_FG_ and M_FW_ groups.

Table S9 The differentially abundant ASVs across the M_C_, M_G_ and M_WG_ groups.

Table S10 The differential colon content metabolites across the M_C_, M_G_ and M_WG_ groups. Table S11 The differentially abundant ASVs between the M_G_ and M_WG_ groups.

Table S12 Primer sequences for RT–qPCR.

